# Detecting Stigmatizing Language in Clinical Notes with Large Language Models for Addiction Care

**DOI:** 10.1101/2025.08.08.25333315

**Authors:** R. Sethi, J. Caskey, Y. Gao, M.M. Churpek, T.A. Miller, A. Mayampurath, E.S. Afshar, M. Afshar, D. Dligach

## Abstract

**RATIONALE:** Recent studies have found that stigmatizing terms can incline physicians to pursue punitive approaches to patient care. The intensive care unit (ICU) contains large volumes of progress notes that may contain stigmatizing language, which could perpetuate negative biases against patients and affect healthcare delivery. Patients with substance use disorders (alcohol, opioid, and non-opioid drugs) are particularly vulnerable to stigma. This study aimed to examine the performance of Large Language Models (LLMs) in the identification of stigmatizing language from ICU progress notes of patients with substance use disorders (SUD).

**METHODS:** Clinical notes were sampled from the Medical Information Mart for Intensive Care (MIMIC)-III, which contains 2,083,180 ICU notes. These 2,083,180 notes were passed into a rule-based labeling approach followed by manual verification for more ambiguous cases. The labeling approach followed the NIH guidelines on stigma in SUD. The labeling process resulted in identifying 38,552 stigmatizing encounters. To design our cohort, we randomly sampled an equivalent amount of non-stigmatizing encounters to create a dataset with 77,104 notes. This cohort was organized into train/development/test datasets (70/15/15). We utilized Meta’s Llama-3 8B Instruct LLM to run the following experiments for stigma detection: (1) prompts with instructions that adhere to the NIH terms (Zero-Shot); (2) prompts with instructions and examples (in-context learning); (3) in-context learning with a selective retrieval system for the NIH terms (Retrieval Augmented Generation-RAG); and (4) supervised fine-tuning (SFT). We also created a baseline model using keyword search. Evaluation was performed on the held-out test set for accuracy, macro F1 score, and error analysis. The LLM-based approaches were prompted to provide their reasoning for label prediction. Additionally, all approaches were evaluated on an external validation dataset from the University of Wisconsin (UW) Health System with 288,130 ICU notes.

**RESULTS:** SFT had the best performance with 97.2% accuracy, followed by in-context learning. The LLMs with in-context learning and SFT provided appropriate reasoning for false positives during human review. Both approaches identified clinical notes with stigmatizing language that were missed during annotation (10/93 false positives for SFT and 22/186 false positives for the in-context learning approach were considered valid after human review). SFT maintained its accuracy at 97.9% on a similarly balanced external validation dataset.

**CONCLUSION:** Our findings demonstrate that LLMs, particularly using SFT and in-context learning, effectively identify stigmatizing language in ICU notes with high accuracy while explaining their reasoning in an asynchronous fashion without needing rigorous and time-intensive manual verification involved in labeling. These models also demonstrated the ability to identify novel stigmatizing language not explicitly in training data nor existing guidelines. This study highlights the potential of LLMs in reducing stigma in clinical documentation, especially for patients with SUD. These LLMs enable identification of stigmatizing language in clinical notes that can perpetuate negative stigma towards patients and encourage rewriting of notes.

## 1. Introduction

Stigmatizing language in electronic health records (EHRs) can hinder effective communication between patients and healthcare providers, influencing clinical decision-making and potentially reinforcing health disparities. The 21st Century Cures Act requires that EHR notes be made available to patients in real-time, online, and at no cost through Open Notes, dramatically increasing the visibility of clinical documentation to both patients and providers [1]. This transparency has heightened the impact of language choice, as terms like “substance abuser” have been shown to provoke more negative provider attitudes and reduce the likelihood of appropriate treatment, compared to more clinically accurate, person-centered alternatives like “substance use disorder” [2]. With the rapid integration of large language models (LLMs) in healthcare settings, the risk of amplifying these biases at scale has become a pressing concern. AI-generated notes and clinical summaries, trained on existing human-authored texts, inherently reflect the biases of their underlying data, potentially perpetuating harmful language if left unchecked [3]. As major EHR vendors are now capable of incorporating LLMs to streamline documentation [4], proactive strategies are essential to prevent the unchecked spread of stigmatizing language in clinical practice [2]. LLMs offer a unique opportunity to drive positive change, serving as tools to reduce stigma, educate providers about the impact of language, and promote documentation practices that build trust and support patient-centered care.

This work explores automated methods for detecting stigmatizing language with respect to addiction within clinical text using LLMs. Our study utilizes the publicly available Medical Information Mart for Intensive Care (MIMIC-III) dataset, which comprises over 2 million intensive care unit (ICU) notes [6]. We hypothesize that LLM-based approaches will outperform simple keyword searches to detect stigmatizing language in accordance with National Institute on Drug Abuse (NIDA) best practices, offering a more accurate and scalable solution for identifying bias in clinical documentation for patients with Addiction Disease. Identification of bias would prompt the clinician to be more aware of the impact of stigma on patient care.

## 2. Related Works

The objective of this study is to design a natural language processing (NLP)-based tool that can classify clinical notes containing stigmatizing language related to substance use. To date, relatively few studies have directly addressed this task. A recent study by Weiner et al. [7] developed a rule-based, closed-source NLP system to detect substance use–related stigmatizing language in clinical notes. Their approach combined a curated list of stigmatizing terms, inspired by the NIDA guidelines [6], with regular expressions and rule-based contextual checks to classify sentences as stigmatizing or non-stigmatizing.

Other studies have focused on detecting stigmatizing language in clinical documentation from various domains, including obstetrics and gynecology notes [8, 9, 10]. These approaches typically used traditional machine learning methods—including decision trees, logistic regression, and support vector machines—combined with features such as term frequency-inverse document frequency (TF-IDF) vectorization of clinical text and patient demographics.

A complementary line of work has investigated sentence-level analysis of stigmatizing language using transformer-based models such as bidirectional encoder representations from transformers (BERT) [11, 12]. These methods first extract potentially stigmatizing sentences using predefined term lists and regular expressions, then classify the contextual use of those terms at the sentence level using transformer models.

In contrast to these prior approaches, our study leverages generative decoder-only LLMs to perform context-aware classification of entire clinical notes. This enables the model to capture both explicit and subtle forms of stigmatizing language without relying solely on predefined term lists or sentence-level parsing.

Our study differs from prior work in several important ways. First, the methods we evaluate are capable of processing entire clinical notes holistically, without requiring any feature engineering, parsing, or pre-selection of potentially stigmatizing sentences. Prior approaches typically rely on keyword-based extraction steps that can miss semantically equivalent but lexically distinct expressions of stigma. Second, unlike sentence-by-sentence evaluation pipelines, which require many independent executions of transformer models, our decoder-only LLM-based approaches can process full clinical notes in a single model execution, leveraging more global context and more efficient computation. Our LLM-based approaches were also found to explain its reasoning behind label predictions, unlike prior approaches.

Finally, our results demonstrate that LLM-based methods can move beyond simple dictionary matching to identify novel, contextually stigmatizing language not explicitly present in predefined guidelines—an essential capability as clinical language continues to evolve.

## 3. Results

### 3.1 Labeled Cohort Demographics and Language Patterns

To create our training and internal testing, we employed a semi-manual approach to identify and label 38,552 clinical notes (from MIMIC-III) containing stigmatizing language and randomly sampled an equivalent number of clinical notes without stigmatizing language. A breakdown of the demographics of the all the MIMC-III data that was split into training, validation, and test sets, provided in **Table 1**. **Table 2** outlines the most frequent stigmatizing terms.

**Table 1.** Data Demographics. Demographics of post-selected and labeled MIMIC-III data [5] (Train, Test, and Validation sets combined), External Validation Full (UW), and External Validation down-sampled/balanced (UW).

| Demographic Characteristic | Selected MIMIC Data (n = 77,104) | External Validation Full (n = 288,130) | External Validation Down-Sampled (n = 4,144) |
| --- | --- | --- | --- |
| Age (yrs), mean (SD) | 56.37 (41.49) | 59 (16) | 55 (15) |
| Sex, n (%) |  |  |  |
| Male | 47,844 (62%) | 176285 (61%) | 2614 (63%) |
| Ethnicity, n (%) |  |  |  |
| Black or African American | 7,298 (9.47%) | 1799 (4.41%) | 208 (5.02%) |
| Pacific Islander or Hawaiian Native | 12 (0.017%) | 67 (0.16%) | 13 (0.31%) |
| White or Caucasian | 51,994 (67.43%) | 36363 (89.04%) | 3641 (87.86%) |
| American Indian or Alaska Native | 76 (0.099%) | 299 (0.73%) | 35 (0.84%) |
| Asian or Mideast Indian | 1,774 (2.30%) | 540 (1.32%) | 42 (1.01%) |
| Hispanic or Latino | 2,319 (3.01%) | 992 (2.43%) | 106 (2.56%) |
| Unknown or Declined | 7,868 (10.20%) | 678 (1.66%) | 88 (2.12%) |
| Non-Hispanic/Latino | 328 (0.46%) | 100 (0.24%) | 11 (0.27%) |

**Table 2.** Top 5 Substance Use-Specific Stigmatizing Language. Top 5 substance use-specific stigmatizing language from post-selected and labeled MIMIC-III data [5] (Train, Test, and Validation sets combined). Note frequency is how many clinical notes this term was identified in. Some of these terms can be from the same note but are considered a unique tally in the frequency count.

| Terms | Note Frequency |
| --- | --- |
| “etoh abuse” | 16,630 (21.56%) |
| “alcohol abuse” | 11,848 (15.36%) |
| “alcoholic cirrhosis” | 4,799 (6.22%) |
| “alcoholism” | 3,925 (5.09%) |
| “polysubstance abuse” | 3,672 (4.76%) |

To create the external validation dataset, we used a data corpus of 288,130 ICU patient clinical notes from UW. This patient data spanned the period from 2009 to 2020, encompassing patients aged 18-86. We also down-sampled the full external validation dataset to select a random convenience sample of 4,144 notes that were evenly split between “yes” and “no” predictions with similar baseline characteristics as the full cohort (**Table 1**). The approaches were evaluated on both the entire external validation dataset and down-sampled dataset. Tables 3, 4 outline the external validations most frequent stigmatizing terms respectively.

**Table 3.** Top 5 Substance Use-Specific Stigmatizing Language in External Dataset Full. Top 5 substance use-specific stigmatizing language from post-selected and labeled External Validation Full. Note frequency is how many clinical notes this term was identified in. Some of these terms can be from the same note but are considered a unique tally in the frequency count.

| Terms | Frequency Full |
| --- | --- |
| “alcohol abuse” | 512 (0.18%) |
| “alcoholic cirrhosis” | 445 (0.15%) |
| “tobacco abuse” | 344 (0.12%) |
| “abuse” | 269 (0.09%) |
| “etoh abuse” | 267 (0.09%) |

**Table 4.** Top 5 Substance Use-Specific Stigmatizing Language in External Dataset Donw-Sampled/Balanced. Top 5 substance use-specific stigmatizing language from post-selected and labeled External Validation Down-Sampled. Note frequency is how many clinical notes this term was identified in. Some of these terms can be from the same note but are considered a unique tally in the frequency count.

| Terms | Frequency Down-sampled |
| --- | --- |
| “alcohol abuse” | 505 (12.19%) |
| “alcoholic cirrhosis” | 438 (10.57%) |
| “tobacco abuse” | 342 (8.25%) |
| “etoh abuse” | 256 (6.18%) |
| “alcoholic” | 253 (6.11%) |

### · **3.1.** Demographics of All Selected MIMIC Data (Train, Validation, Test Sets)

This study was reviewed by the University of Wisconsin-Madison Institutional Review Board (IRB; 2023-1252) and determined to be exempt from human subjects research. The IRB approved the study with a waiver of informed consent.

#### Experiments

Several approaches were experimented with, including a baseline keyword method and several LLM-based strategies for detecting stigmatizing language in addiction care. The baseline method relied on keyword searches using a custom dictionary of stigmatizing language-related terms, compiled from established guidelines on reducing bias in clinical communication for addiction care [6,13]. For the LLM-based approaches, we employed open models such as the Meta-Llama-3-8B-Instruct model [14] using the HuggingFace transformers library [15] and PyTorch [16]. We explored multiple configurations: simple zero-shot prompting, in-context generation and retrieval-augmented generation (RAG) using additional context from stigmatizing language guidelines, and supervised fine-tuning (SFT) to adapt the model specifically to our task. These configurations were designed to capture both explicit and context-dependent instances of stigmatizing language, offering a more flexible and context-aware approach than traditional keyword matching. A summary of these methods is provided in **Figure 1**.

**Figure 1.**
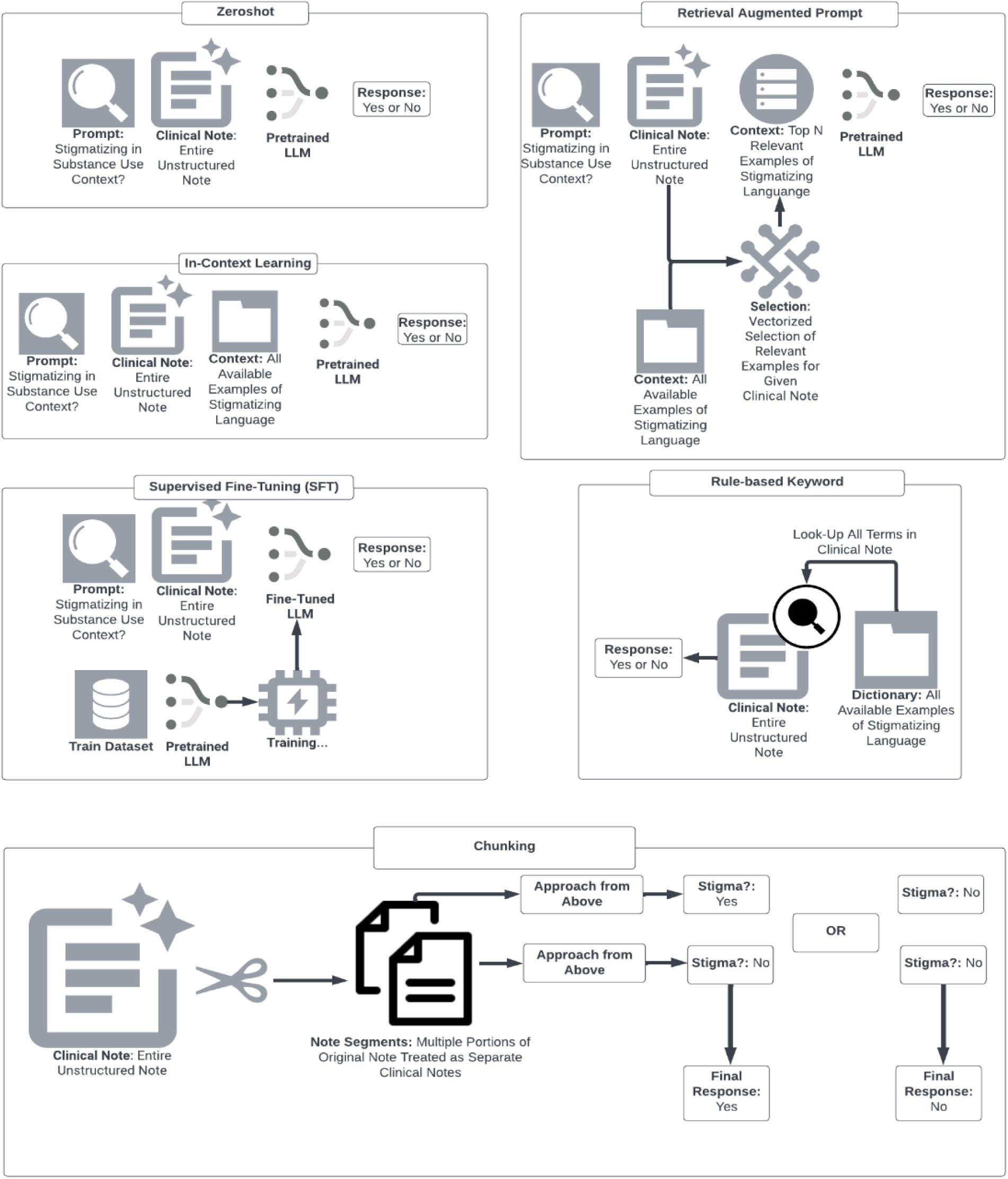
Study Design of Experiments for Stigma Detection. Zero-shot uses a simple prompt that asks an LLM to evaluate a clinical note for stigmatizing language in a substance use context, without any additional context or explanation. In-context learning is a modified version of zero-shot learning, where the prompt is enriched with guidelines on what constitutes stigmatizing language. Retrieval Augmented Generation/Prompting (RAG) modifies the original prompt with guidelines, except that the top n guidelines are retrieved using a selection method between the clinical note and guideline entry. Supervised Fine-Tuning (SFT) follows the same prompt structure as zero-shot, except that the model is fine-tuned to the task. Rule-based keyword performs a text look-up for a list of commonly used stigmatizing terms from NIDA guidelines [6]. Chunking is a technique that is integrated with any of the approaches explained above, where clinical notes are broken into segments and each segment is processed by an approach independently. After those chunks are processed, the final output depends on the outcome of each segment (i.e., if any segment is classified as stigmatizing, the entire note is labeled as positive for stigma).

We evaluated our models on a fully held-out test set, as well as on a more challenging subset of the test set. Every note in this challenging subset contains terms in a list of terms inspired by NIDA guidelines [6]. Upon manual review of the surrounding context around the flagged term, some of these notes were labeled as non-stigmatizing despite their potential for misinterpretation. This latter evaluation was designed to test the models’ ability to distinguish stigmatizing language from contextually appropriate use (i.e. patient has a ‘junky’ cough), reflecting the nuanced nature of real-world clinical documentation. To evaluate model performance, we employed accuracy and F1 scores for the automated identification of stigmatizing language with 1000-iteration bootstrapped 95% Confidence Intervals (95% CI). Detailed manual error analysis was also performed on cases missed by the LLM. To assess the generalizability of the approaches tested, we also evaluated the approaches on an external validation dataset from UW. The external validation dataset was utilized in both native prevalence and down-sampled balanced prevalence of stigmatizing labels. This was to evaluate the approaches on unseen data with true prevalence of stigmatizing labels (entire dataset) and the down-sampled version to mimic the same distribution labels utilized on the MIMIC-based test set.

#### Findings

The results of the performance evaluation on the full held-out test set are presented in **Table 5**. The SFT approach achieved the highest performance for identifying substance use-related stigmatizing language, with a bootstrapped macro-averaged F1 score of 0.970 (95% CI: 0.970 - 0.973), followed by the in-context model, which achieved a score of 0.890 (95% CI: 0.889 - 0.895) (**Table 5**). In contrast, the baseline keyword search approach performed worse, with a macro-averaged F1 score of 0.680 (95% CI: 0.678 - 0.687). Notably, the in-context learning approach, which leveraged task-specific context during inference, approached the performance of the SFT model despite requiring significantly less task-specific training data, highlighting its potential as a more alternative parsimonious model.

**Table 5.** Model Performance on the Full Held-Out Test Set. Bootstrapped (n = 1,000) performance on the complete held-out test set (11,586 clinical notes). Each approach reflects a distinct prompting or fine-tuning strategy for identifying stigmatizing language. Results are reported as mean accuracy and macro F1 scores with 95% bootstrapped confidence intervals.

| Approach | Accuracy | Macro F1 |
| --- | --- | --- |
| Supervised-Fine Tuning (SFT) | 0.972 (0.970 - 0.973) | 0.972 (0.970 - 0.973) |
| In-context | 0.892 (0.889 - 0.895) | 0.892 (0.889 - 0.895) |
| Retrieval Augmented Prompt (RAG) | 0.889 (0.886 - 0.892) | 0.889 (0.886 - 0.892) |
| Zero-Shot Prompt | 0.834 (0.831 - 0.838) | 0.834 (0.831 - 0.838) |
| Keyword | 0.683 (0.678 - 0.687) | 0.678 (0.673 - 0.682) |

Additionally, RAG performed similarly to the In-Context approach on the full test set (**Table 5**), whereas Zero-shot lagged with an approximately 0.05 drop in F1 score (**Table 5**). However, all these approaches outperformed the baseline keyword approach with gains of at least 0.17 F1 score (**Table 5**).

The performance evaluation on the challenging subset of the test set is summarized in **Table 6**. The SFT approach achieved the highest macro-averaged F1 score of 0.896 (95% CI: 0.890 - 0.902), outperforming the in-context learning approach, which reached 0.690 (95% CI: 0.683 - 0.698). Notably, the SFT approach exhibited less performance degradation when transitioning from the full test set to this more challenging subset, with its F1 score decreasing by just 0.078, compared to a 0.202 decrease for the in-context approach. In contrast, the baseline keyword search method, which lacks the capacity to interpret context, performed worse, with a macro-averaged F1 score of 0.340 (95% CI: 0.337 - 0.344), compared to the SFT model. These results highlight the advantage of context-aware LLM-based methods in distinguishing between stigmatizing language in contextually appropriate use, even in challenging scenarios.

**Table 6.** Model Performance on a Challenging Subset of the Test Set. This challenging subset contains stigmatizing terms which could make the clinical note stigmatizing or non-stigmatizing depending on the context it’s surrounded with. Bootstrapped (n = 1,000) performance on a subset of 6,889 clinical notes from the held-out test set, each containing one or more stigmatizing terms. Labels were assigned based on manual review of contextual usage by addiction care expert (ESA), reflecting whether the term was used in a genuinely stigmatizing or non-stigmatizing manner. Results are reported as mean accuracy and macro F1 scores with 95% bootstrapped confidence intervals.

| Approach | Accuracy | Macro F1 |
| --- | --- | --- |
| Supervised-Fine Tuning (SFT) | 0.961 (0.959 - 0.964) | 0.896 (0.890 - 0.902) |
| In-context | 0.833 (0.828 - 0.837) | 0.690 (0.683 - 0.698) |
| Retrieval Augmented Prompt (RAG) | 0.743 (0.711-0.775) | 0.652 (0.614 - 0.690) |
| Zero-Shot | 0.745 (0.740 - 0.750) | 0.609 (0.603 - 0.616) |
| Keyword | 0.478 (0.472 - 0.484) | 0.340 (0.337 - 0.344) |

The best-performing approach (SFT) and the baseline keyword method were externally validated on the EHR cohort from UW. We evaluated both approaches on the external dataset at its native label prevalence, which comprised 288,130 notes, of which only 2,072 (0.72%) were labeled as stigmatizing by our semi-manual annotation pipeline. As expected, performance declined in this more imbalanced, real-world scenario. The SFT model achieved a macro F1 score of 0.759 (95% CI: 0.747-0.772), while the keyword baseline continued to have lower performance at 0.699 (95% CI: 0.682-0.715) (Table 7). Despite this drop, the SFT model maintained superior performance over the baseline, demonstrating its ability to generalize beyond the training distribution and handle low-prevalence settings more effectively. We also evaluated both approaches on a balanced subset of the full external dataset. This balanced dataset contained 4,144 clinical notes, evenly split between stigmatizing and non-stigmatizing labels to mirror the data distribution used during model training. On this balanced set, the SFT model demonstrated robust generalization, achieving a macro F1 score of 0.978 (95% CI: 0.974-0.983) and outperforming the keyword baseline, which achieved a macro F1 score of 0.849 (95% CI: 0.838-0.860) (**Table 8**).

**Table 7.** External Validation on the Full External Dataset at Native Label Prevalence. Bootstrapped (n = 1,000) performance on the complete external dataset (288,130 clinical notes), in which stigmatizing language was present in 2,072 notes (0.72%). Results reflect model performance in a real-world, highly imbalanced setting without artificial rebalancing of label distribution.

| Approach | Accuracy | Macro F1 |
| --- | --- | --- |
| Supervised-Fine Tuning (SFT) | 0.996 (0.995-0.996) | 0.759 (0.747-0.772) |
| Keyword | 0.996 (0.995-0.996) | 0.699 (0.682-0.715) |

**Table 8.**
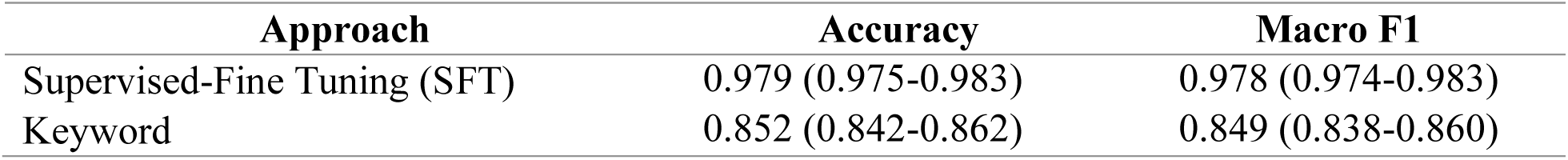
External validation on a Down-Sampled Balanced Subset of the External Dataset. Bootstrapped (n = 1,000) performance on a down-sampled subset of 4,144 clinical notes from the external dataset, evenly balanced between notes labeled as containing stigmatizing language and those labeled as non-stigmatizing (2,072 each). This evaluation mirrors the balanced distribution used during model training and enables direct comparison of model performance across approaches.

| Approach | Accuracy | Macro F1 |
| --- | --- | --- |
| Supervised-Fine Tuning (SFT) | 0.979 (0.975-0.983) | 0.978 (0.974-0.983) |
| Keyword | 0.852 (0.842-0.862) | 0.849 (0.838-0.860) |

#### Error Analysis

To further assess the robustness of LLM-based approaches in identifying stigmatizing language, we conducted an LLM-assisted human-in-the-loop error analysis, focusing on false positive instances—clinical notes that the models labeled as stigmatizing but were annotated as non-stigmatizing. Both the SFT and in-context models were included in this analysis. For each false positive, the models were prompted a second time to explain their reasoning, with the in-context approach receiving the same context used during the initial classification. These explanations were then manually reviewed by an addiction care expert (ESA) to determine whether the original human annotations might have missed genuinely stigmatizing language. The SFT approach correctly identified 10 clinical notes as genuinely stigmatizing upon this second review, while the in-context approach identified 22 such instances (**Table 9**). A comparative analysis of the specific phrases identified by each model, including terms detected in isolation and conjunction, is presented in **Figure 2**. Notably, many of these terms were not explicitly included in the NIDA guidelines [6], highlighting the potential for LLMs to detect contextually stigmatizing language that extends beyond their original training data.

**Table 9.** Error Analysis Results. Results from error analysis evaluating false positives. Unreadable: the model did not coherently explain its reasoning when prompted. Decision switch: the model did not agree with its original answer to the prompt. Unrelated to substance use: the model considered the note stigmatizing in a non-substance use context. Valid: the model identified stigmatizing language regarding substance use.

| Approach | Unreadable | Decision Switch | Too Sensitive or Unrelated to Substance Use | Valid | Total False Positives |
| --- | --- | --- | --- | --- | --- |
| Supervised-Fine Tuning (SFT) | 11 | 45 | 27 | 10 | 93 |
| In-context | 11 | 1 | 152 | 22 | 186 |

**Figure 2.**
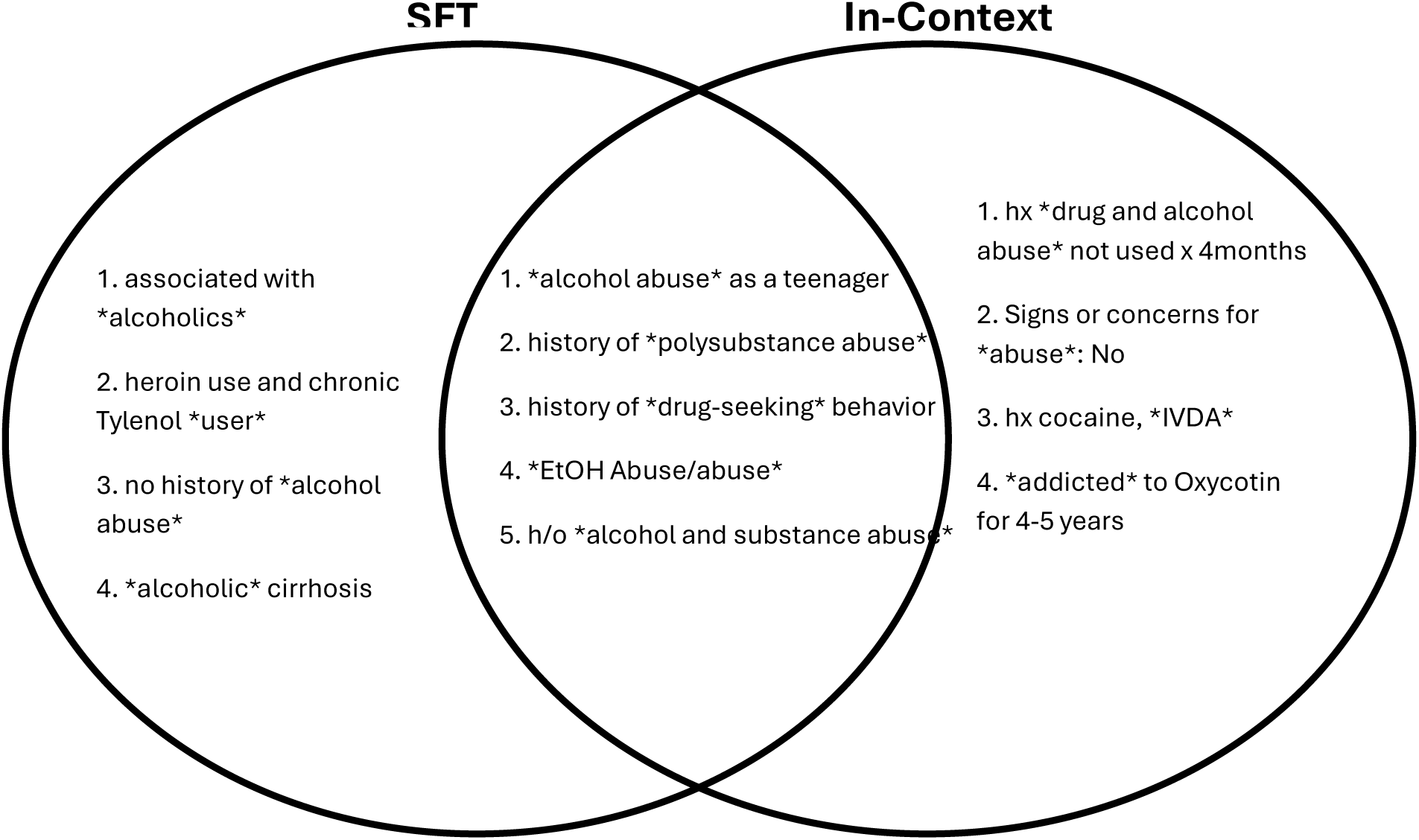
Examples of Previously Unidentified Substance Use-Related Stigmatizing Language. These are examples of substance use-related stigmatizing language from clinical notes that were labeled originally as non-stigmatizing. These examples are isolated from false positives made by SFT and in-context approaches. These false positives were reviewed by an addiction care specialist (ESA) to determine whether the approaches were correct to label these phrases as stigmatizing. Of note, these examples are ones that were not present in the training set nor mentioned in the NIDA guidelines for substance use-related stigmatizing language. The segments in the middle are language identified by both approaches, while the flanking segments are those identified solely by the approach under which they appear. Abbreviations from the examples above: Intravenous Drug Abuse (IVDA), ethanol/alcohol (EtOH), h/o (history of), hx (history). Stigmatizing terms are surrounded by ‘*’.

Another noteworthy finding from the error analysis was the frequency with which the models revised their initial classifications when prompted to provide reasoning. The SFT approach demonstrated significantly more decision switching, with approximately 50% of its original classifications being adjusted upon second review, compared to less than 1% for the in-context approach (**Table 9**). However, expert review revealed that roughly 90% of these false positive instances were indeed genuinely non-stigmatizing, suggesting that the greater consistency observed in the in-context model often reflected a rigidity in its decision-making rather than superior accuracy. This finding underscores a key advantage of the SFT approach: despite being fine-tuned for the specific task of identifying stigmatizing language, it retains flexibility in responding to varied prompts, potentially allowing for more contextually accurate classifications when reconsidering ambiguous cases.

#### Computational considerations

In terms of processing efficiency, the RAG approach had the longest average inference time, requiring 46.46 seconds per note. This extended latency reflects the overhead associated with first identifying the most relevant context entries before generating a response. In contrast, the baseline keyword approach was the fastest, with an average inference time of less than 0.001 seconds per note, owing to its straightforward text-matching design. The SFT approach, while significantly faster than RAG, required a substantial up-front training investment, including approximately 6 hours of fine-tuning on 8 parallel NVIDIA A6000 Graphical Processing Units (GPUs), with an effective batch size of 64 for 3 epochs. Detailed inference speed comparisons for each approach are provided in **Table 10**.

**Table 10.** Processing and Training Time. Average inference time per input was averaged across all evaluation data (excluding external validation). Training time was only relevant to SFT.

| Approach | Average Inference Time per Input (s) | Train Time (hrs) |
| --- | --- | --- |
| **Supervised-Fine Tuning (SFT)** | 4.80 | 5.90 |
| **In-Context Prompt** | 3.37 | n/a |
| **Retrieval Augmented Prompt (RAG)** | 46.46 | n/a |
| **Zero-Shot** | 1.57 | n/a |
| **Keyword** | <0.001 | n/a |

## 4. Discussion

This study demonstrates that the LLM-based approaches consistently yielded superior performance in detecting stigmatizing language within substance use-related clinical notes compared to keyword search methods. The SFT model performed the best, achieving the highest precision and context sensitivity, a critical requirement for this task. However, the in-context learning approach, despite lacking the task-specific training of the SFT model, achieved competitive performance, highlighting its potential as a more resource-efficient alternative.

RAG and zero-shot approaches served as additional approaches that did not require fine-tuning, similar to the in-context approach. These approaches outperformed the baseline approach, demonstrating the advantage of utilizing LLMs over simple keyword search algorithms for stigmatizing language. However, in-context performed the best from these approaches on the challenging test subset and warranted further comparisons with the SFT approach to investigate the benefits of additional fine-tuning on this task.

Both the SFT and in-context approaches demonstrated the ability to identify novel, contextually stigmatizing language that was not explicitly included in their training data or predefined guidelines. For instance, the SFT model accurately flagged terms like “drug-seeking behavior” and “alcoholic cirrhosis,” which were not part of the NIDA guidelines [6] used to annotate the training dataset. This suggests that these models can extend beyond the specific terms they were trained on, potentially identifying emerging stigmatizing language as clinical documentation evolves, unlike simple baseline keyword searches.

Results from external validation demonstrated that the SFT approach generalized well to unseen data. As expected, the SFT model exhibited a performance decline in external validation when evaluated on the external dataset at its native prevalence of stigmatizing language, where non-stigmatizing notes vastly outnumbered stigmatizing ones. This drop was likely attributable to the model’s heightened sensitivity, which was calibrated during training on a balanced dataset, potentially leading to increased false positives in a highly imbalanced real-world setting. However, the SFT model continued to outperform the baseline keyword approach in this scenario. Importantly, this performance drop could likely be mitigated in practice with domain adaptation on data reflecting the native label distribution (though we did not perform this experiment here due to computational constraints associated with training on a much larger, imbalanced dataset and lack of training appropriate representation for stigmatizing terms in the imbalanced set). Notably, the SFT model retained excellent performance on the balanced subset of the external dataset, mirroring the distribution used during training, further validating the robustness of the fine-tuning strategy.

Despite this advantage, the SFT model also demonstrated a notable trade-off in flexibility. During error analysis, the SFT model was more responsive to varied prompts, effectively reconsidering its classifications upon second review. However, this adaptability came at the cost of consistency, with the SFT model reversing its initial decisions more often than the in-context approach. Given that most of these reversals were ultimately found to be unnecessary upon expert review, this finding suggests that the SFT model’s flexibility might increase the risk of false-positive identification, particularly in ambiguous cases.

In contrast, the in-context learning approach, which relies on prompting without additional task-specific training, maintained more consistent decision-making during the error analysis phase, revising fewer initial classifications. This consistency may be advantageous in clinical applications where reliability is critical, although further evaluation across a broader range of clinical contexts is necessary to confirm this finding.

Nevertheless, the SFT model demonstrated a superior ability to account for context, as evidenced by its more stable performance on a challenging subset of test notes. This subset, which included terms that may or may not be stigmatizing depending on the context, revealed a sharper decline in performance for the in-context approach, reinforcing the SFT model’s robustness in distinguishing genuinely stigmatizing language from contextually appropriate use.

Overall, while the in-context learning approach offers a promising, low-cost alternative for rapidly aligning LLMs to evolving guidelines, the SFT approach was the more precise and context-aware option, particularly in high-stakes medical applications

## 5. Conclusion

SFT demonstrated the highest accuracy in detecting stigmatizing language in clinical notes, particularly within substance use contexts, offering superior context sensitivity and precision. However, the in-context learning approach achieved competitive results without the resource-intensive requirements of custom training, highlighting its potential for rapid deployment in evolving clinical settings. Both methods identified stigmatizing language beyond their training data, reflecting the broader linguistic knowledge of large language models. Notably, while the SFT model exhibited greater flexibility in reconsidering ambiguous cases, it also showed a higher tendency for decision reversals, underscoring the trade-offs between adaptability and consistency in real-world applications. Future efforts should explore hybrid approaches that leverage the strengths of both fine-tuned and in-context methods to enhance the accuracy, interpretability, and scalability of stigma detection in clinical documentation.

## 6. Limitations and Future Works

All experiments were performed utilizing LLMs with 8B parameters or less due to memory constraints. Furthermore, a few clinical notes exceeding the input token restrictions prevent accurate predictions for those notes. Finally, although our dataset contained over 70,000 examples for training and evaluation, only around 4,000 of those examples were clinical notes that contained stigmatizing terms and were considered non-stigmatizing. Despite its high performance in internal and external validation, the SFT model trained could have benefited from more of such examples.

Future work should explore hybrid strategies that combine the contextual strength of fine-tuning with the consistency of in-context methods, potentially improving both accuracy and interpretability in real-world deployments. Naturally, the next step involves building on findings from the identification of stigmatizing language and designing a system for correcting such language without modifying the meaning of the original text.

## 7. Methods

### 7.1 Dataset Generation

We used the MIMIC-III, a publicly available, de-identified database comprising over 2 million clinical notes from ICU stays at Beth Israel Deaconess Medical Center between 2001 and 2012 [5]. MIMIC-III is widely utilized in medical informatics research due to its scale, granularity, and real-world clinical context.

To identify candidate records for our analysis, we leveraged a set of substance use–related stigmatizing terms defined by NIDA [6]. Using this term list, we performed a keyword search across 2,083,180 progress notes within MIMIC-III [5], extracting 42,641 notes that contained at least one potentially stigmatizing term (e.g., “addict,” “drunk”). 4,089 of these notes are considered non-stigmatizing despite containing terms that are potentially stigmatizing. 34,463 notes were sampled from the progress notes no containing any potentially stigmatizing terms to create an evenly balanced dataset of both labels.

In the MIMIC-III [5] corpus, clinical notes often exist in clusters corresponding to a single patient encounter (e.g., iterative modifications to the same note). To prevent data leakage across training, validation, and test sets, we first grouped notes by encounter ID, patient ID, and caregiver ID. These groups were then assigned to splits using a stratified 70-15-15 (train-validation-test) strategy, ensuring that no encounter was represented in more than one split and limit data leakage. Breakdown of the dataset in **Table 11 and 12**.

**Table 11.** Breakdown of Entire Dataset. Label and average token length statistics for post-processed and selected MIMIC-III [5] data.

| **Total Notes** | **Notes Containing Stigmatizing Language** | **Notes Containing Potentially Stigmatizing Terms but Not Used in Stigmatizing Context** | **Avg Token Length** |
| --- | --- | --- | --- |
| 77,104 | 38,552 | 4,089 | 1250.10 |

**Table 12.**
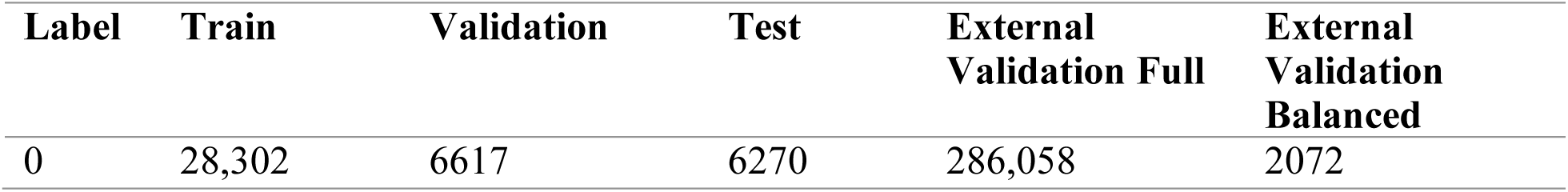
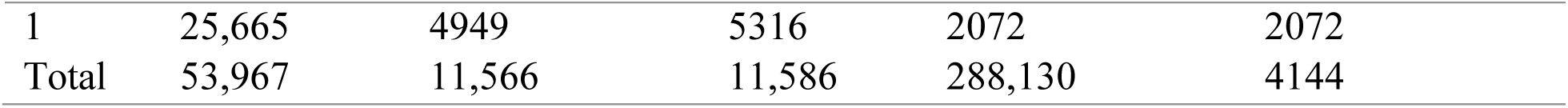
Breakdown of Labels for Each Dataset Split. Breakdown of labels for train, validation, and test sets from post-processed and selected MIMIC-III [5] data and external validation sets from University of Wisconsin-Madison Hospital. External validation breakdown is provided for the native prevalence of the label (no selection, simply recording the true frequency of stigmatizing language) and a randomly balanced set.

To create the external validation dataset, we used a data corpus of 288,130 patient clinical notes from UW. This patient data spanned 2009-2020 from patient ages 18-86, and the external validation dataset only included notes that were from patients in the ICU. Since this dataset included PHI, all analyses with the external validation dataset took place in a HIPAA-secure computing environment. For the balanced external validation dataset, we down-sampled the full external validation dataset to 4,144 notes that were evenly split between “yes” and “no” labels. Approaches were evaluated on both the full and balanced external validation datasets. Breakdown of the dataset in **Table 12**.

### 7.2. Reference Labels and Primary Outcome

The primary outcome of this study was the automated identification of stigmatizing language in clinical documentation as a classification task. Reference labels were generated through a semi-manual annotation process designed to distinguish stigmatizing language from non-stigmatizing or contextually appropriate usage.

From the initial subset of 42,641 notes containing stigmatizing terms, each note was semi-manually reviewed and labeled as either stigmatizing or non-stigmatizing based on a set of predefined, context-aware criteria inspired by NIDA [6]. These criteria accounted for the presence of quotation marks, speaker attribution, and linguistic context. For example, the phrase *“he feels ‘stinking drunk’ after…”* includes the term “drunk,” but was not labeled as stigmatizing because the language was attributed to the patient language, not the provider. There was some manual review involved for inputs that did not meet all criteria outlined in Figure 3. This review was performed by an addiction specialist (ESA) who also provides training to clinicians on appropriate use of language and stigma.

**Figure 3.**
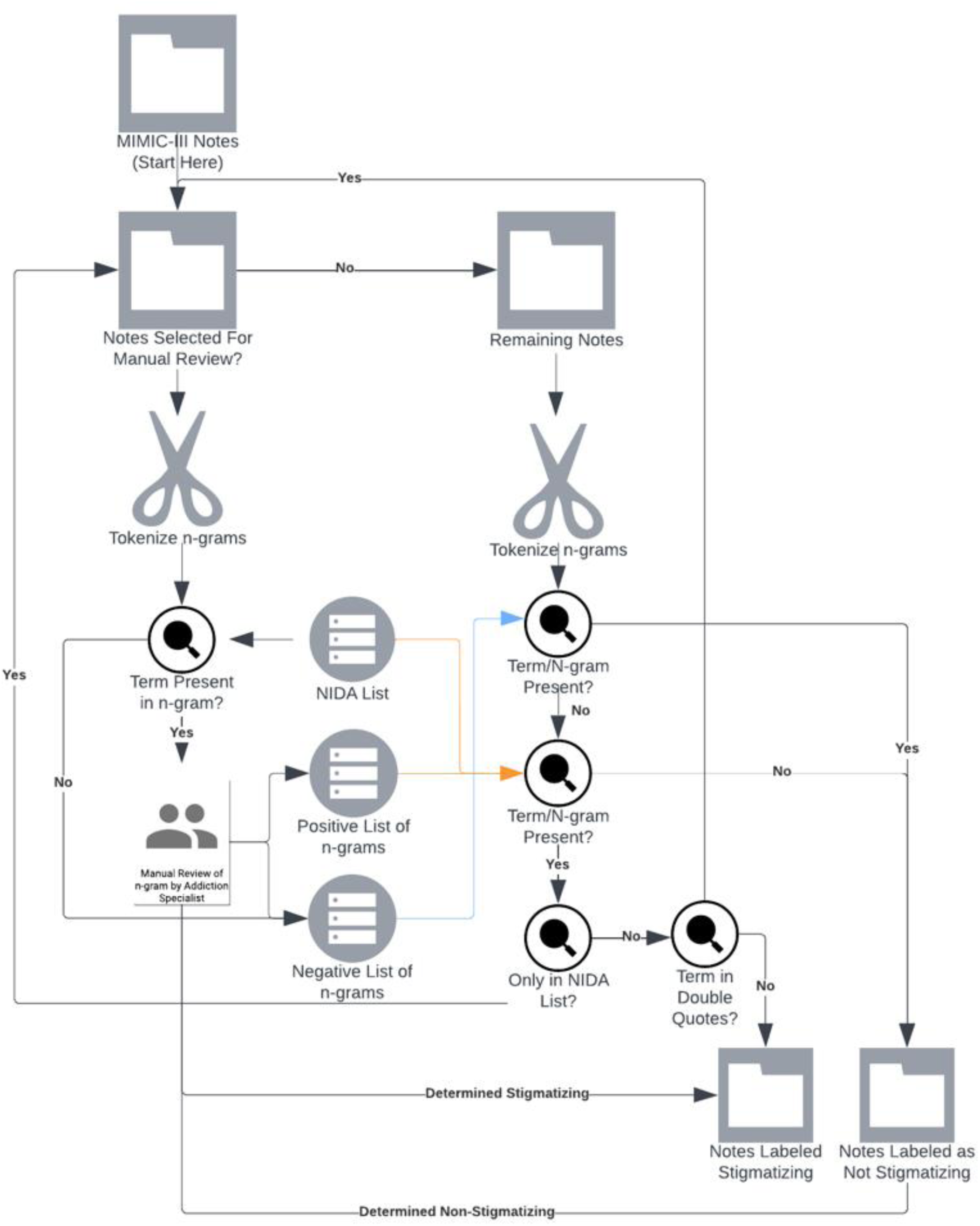
Dataset Labeling Workflow. Annotation required semi-manual review of clinical notes from MIMIC-III [5] and UW External Validation Datasets. An adapted term list was generated using NIDA guidelines [6] and used to identify a preliminary list of stigmatizing notes. This was followed by a set of rules to determine truly stigmatizing terms. Those notes that were still not confirmed to be stigmatizing were tagged for manual review by an addiction specialist (ESA).

This review process resulted in 38,552 notes labeled stigmatizing and 4,089 notes labeled non-stigmatizing from manual review. To ensure class balance and enable more robust model training, we randomly sampled an additional 34,478 notes from the remainder of the MIMIC-III [5] dataset that did not contain any stigmatizing terms or language. These notes were labeled as non-stigmatizing. Breakdown on the notes is summarized in **Table 11**.

The final dataset comprised three distinct categories: (1) stigmatizing, i.e. notes containing stigmatizing terms used in a stigmatizing context; (2) contextually non-stigmatizing, i.e. notes containing stigmatizing terms not used in a stigmatizing context; (3) non-stigmatizing, i.e. notes without any stigmatizing terms or language.

To ensure balanced representation across all stages of model development, we maintained consistent distributions of these categories across the training, validation, and test sets (70-15-15). An overview of the dataset development process is outlined in **Figure 3**. We followed the transparent reporting of a multi-variable model for individual prognosis or diagnosis (TRIPOD)-LLM guidelines, and the accompanying checklist is available in the Supplementary Table 1 (S1).

The same pipeline for labeling the MIMIC-based dataset was applied to the external validation dataset for labeling (minus dataset splits as external validation was treated as a test set).

### 7.3 Experimental Design

To evaluate the effectiveness of various strategies for detecting stigmatizing language in clinical text, we implemented and compared five approaches: a keyword-based baseline and four configurations leveraging an LLM **(Figure 1).** All LLM-based methods utilized the Meta-Llama-3-8B-Instruct model [14], an open-source, instruction-tuned LLM released by Meta. This model supports a range of inference paradigms, including zero-shot prompting, RAG, in-context learning, and SFT.

#### Keyword-Based Baseline

The keyword-based method served as a baseline. Clinical notes were tokenized using whitespace, and each token was compared to a predefined list of stigmatizing terms derived from the NIDA Guidelines [6]. If a note contained any matching term (regardless of context), it was labeled as stigmatizing. This approach does not account for quotation usage or speaker attribution; therefore, it cannot differentiate between stigmatizing intent and neutral or patient-attributed mentions.

#### Zero-Shot Prompting

The zero-shot approach involved prompting the LLM with a direct question: *“Does this note contain stigmatizing language in a substance use context?”* The prompt was designed to elicit binary (“yes” or “no”) responses and was optimized using the validation set. This configuration provided no additional context or examples. Model outputs were converted to binary labels to facilitate evaluation. The sample prompt format is shown in **Figure 4**.

**Figure 4.**
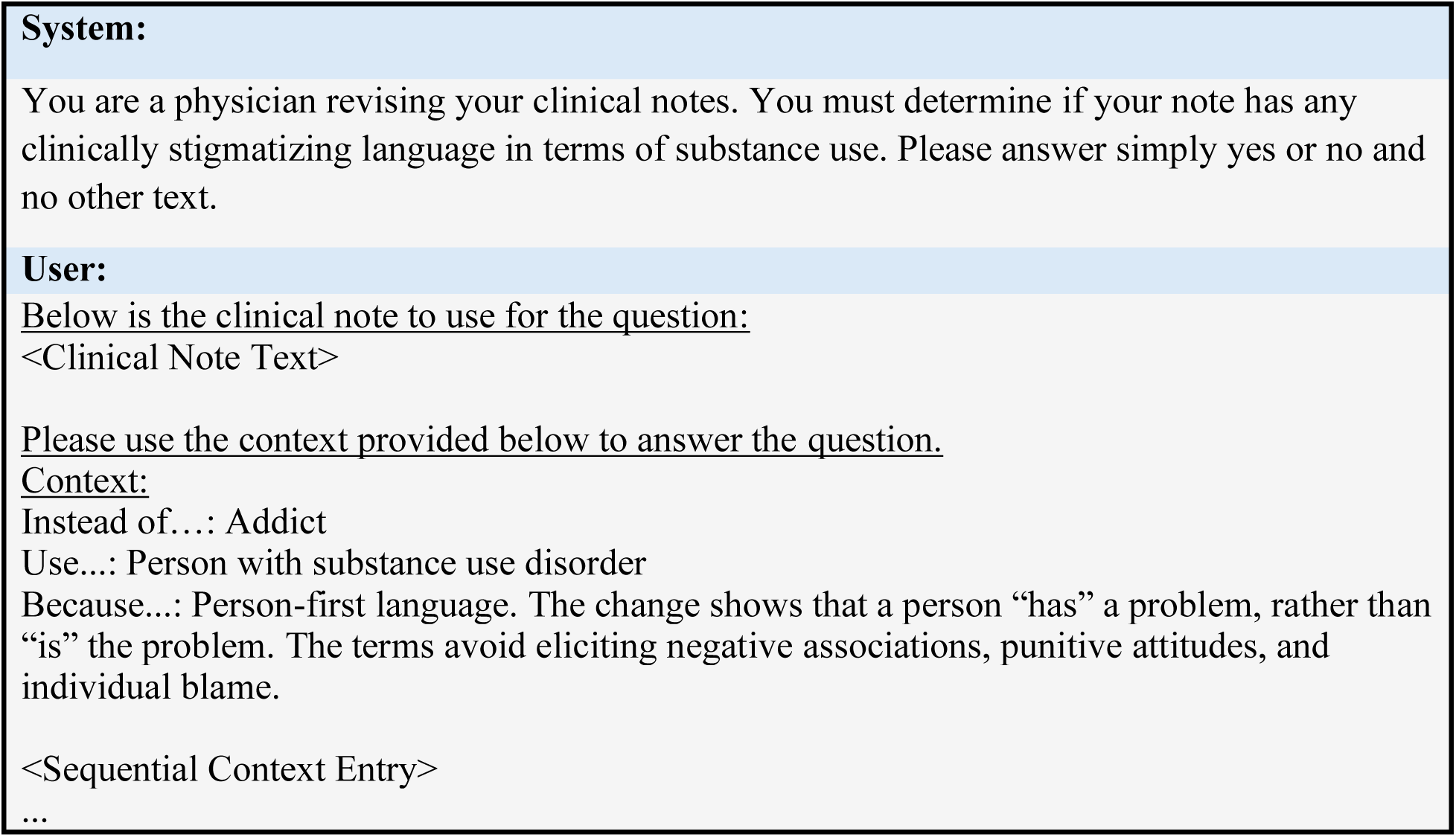
General Structure of Prompt. When applying this prompt to the RAG approach, there are n context entries. When applying the in-context approach, all available context entries are provided. Finally, when applying to the Zero-Shot or SFT approach, there is no context subfield in the user portion of the prompt. System prompts prime the LLM to answer user prompts following certain directions. Hence the system prompt gives the LLM a scenario and an output format.

#### Retrieval-Augmented Generation (RAG)

The RAG configuration enhanced the base prompt with relevant contextual information drawn from external guideline sources. These sources included structured examples and definitions of stigmatizing language [6, 13]. Each guideline entry was encoded into an embedding using the Meta-Llama-3-8B-Instruct model [14]. Input clinical notes were similarly embedded, and cosine similarity was computed to identify the top-n most relevant entries. These were appended to the prompt as contextual guidance. The number of retrieved entries, n, was treated as a hyperparameter and optimized using the validation set. This approach aimed to strike a balance between contextual specificity and prompt length. Prompt formatting is illustrated in **Figure 4**.

#### In-Context Learning

This configuration provided the full set of guideline-derived context entries alongside the clinical note, without filtering based on similarity in the RAG approach. While less selective than RAG, this approach ensured comprehensive access to guidance examples. The same source materials were used as in the RAG setup.

#### Supervised Fine-Tuning (SFT)

In the SFT configuration, the Meta-Llama-3-8B-Instruct [14] model was fine-tuned on the labeled dataset to specialize it for the task of stigmatizing language detection. The input format during inference matched that of the zero-shot configuration. Fine-tuning was performed using the LLaMA Factory [17] implementation of Parameter-Efficient Fine-Tuning (PEFT) via Low-Rank Adaptation (LoRA).

Additionally, the DeepSpeed integration within LLaMA Factory [17] was employed to accelerate training and optimize memory usage.

### 7.4. Implementation Details

This section outlines key low-level design choices and optimization strategies implemented across our LLM-based methods, including the selection and structuring of external guidelines for prompt augmentation, input chunking for handling long notes, and the hyperparameter tuning process used to optimize each configuration. Meta-Llama-3-8B-Instruct [14] was implemented for all LLM-based approaches. The final use for inference and training on 03/2025.

#### 7.4.1. External Guidelines for Context Augmentation

For both the RAG and In-Context approaches, we incorporated external clinical communication guidelines to provide the model with explicit definitions and examples of stigmatizing language. These guidelines were presented as structured tables in the source literature, with each entry describing a category of stigmatizing language alongside representative examples. Two distinct resources were used for context:

1. NIDA Stigmatizing Language Guidelines [6], which provide substance use–specific recommendations and include 14 table entries.
2. A Systematic Review of Stigmatizing Language in Healthcare [13], which offers broader guidance encompassing both substance use–related and general stigmatizing language. This resource contains 30 entries.

Each entry was processed into an embedding vector using the Meta-Llama-3-8B-Instruct [14] model via Hugging Face’s feature-extraction pipeline [15]. These embeddings were stored and used to compute similarity scores with input clinical notes for the RAG method. For both RAG and In-Context prompting, the source of guideline entries (either [6] or [13]) and the number of entries used were treated as tunable hyperparameters.

#### 7.4.2. Input Chunking

Due to token limitations inherent in transformer-based LLMs and the potentially long length of clinical progress notes, we implemented a chunking strategy for the Zero-Shot, RAG, and Full-Context configurations. Each note was divided into overlapping segments of length N tokens, with an overlap of M tokens between adjacent chunks. Each chunk was processed independently using the standard prompting schema (as shown in **Figure 4**). If any chunk was classified as containing stigmatizing language, the entire note was labeled as stigmatizing.

Both the chunk size, N, and overlap, M, were treated as hyperparameters. Chunking was evaluated against non-chunked versions of each method to assess performance trade-offs, particularly in the context of long clinical notes.

#### 7.4.3. Hyperparameter Optimization

Hyperparameter tuning was performed using the validation set for each model configuration. The following hyperparameters were explored:

Not all hyperparameters were applicable to all methods; a full breakdown is provided in **Table 13**. For each approach, every combination of valid hyperparameters was evaluated on the validation set, and the best-performing configuration was selected and used for final model testing. The final selected hyperparameters are defined in **Table 14**. Hyperparameters used for fine-tuning are provided in **Table 15**.

**Table 13.** All Possible Hyperparameters for Approaches. Hyperparameters explored for approaches investigated. RAG similarity metric was the metric utilized to determine the similarity between a clinical note and guideline entry and is only relevant for the RAG approach. Number of context entries is how many guideline entries are included in the prompt of the RAG approach. Context source is what resource the guideline entries are extracted from and relevant for RAG and In-context approaches. Finally, chunking segments of clinical notes. Hyperparameters for chunking include the size of the chunks and overlap between each chunk.

| Approach Hyperparameter | Values |
| --- | --- |
| RAG Similarity Metrics (RAG only) | Cosine, Dot Product |
| Number of Context Entries (RAG only) | 1, 3, 5 entries |
| Context Source (RAG and In-Context) | [6] NIDA Guidelines, [13] Systematic Review |
| Chunking Parameters |  |
| Chunk Size | None, 200, 1000 tokens |
| Overlap | None, 20, 100 tokens |

**Table 14.** Best Performing Hyperparameters for All Approaches. These hyperparameter values were selected for using the results from the validation dataset where all hyperparameter sets were scored on this subset of the MIMIC-III [5] data.

| Approach | Chunk Size (Tokens) | Chunk Overlap (Tokens) | Scoring | Num. Context Entries | Context Source |
| --- | --- | --- | --- | --- | --- |
| Supervised-Fine Tuning (SFT) | N/A | N/A | N/A | N/A | N/A |
| In-context | 1000 | 100 | N/A | N/A | [6] |
| Retrieval Augmented Prompt (RAG) | 1000 | 100 | dot | 5 | [13] |
| Zero-Shot Prompt | 1000 | 100 | N/A | N/A | N/A |
| Manual | N/A | N/A | N/A | N/A | N/A |

**Table 15.**
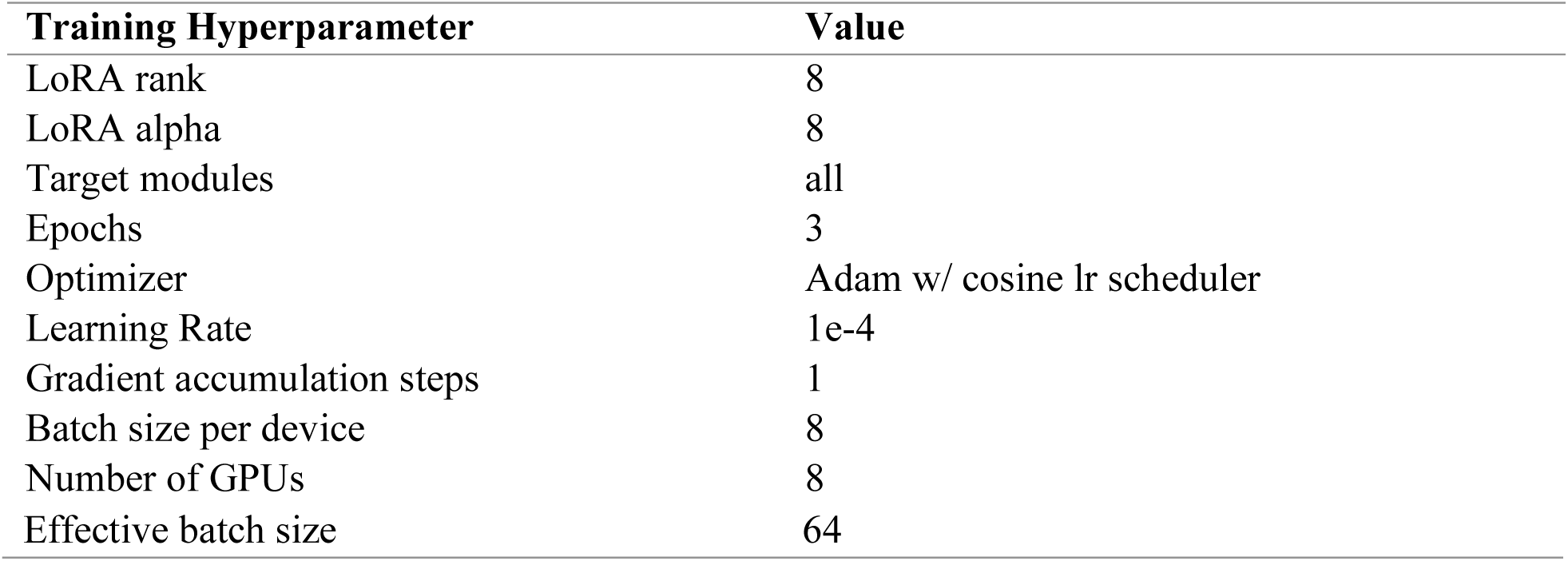
Hyperparameters for Supervised Fine-Tuning (SFT) LLM Approach. These hyperparameters were selected to optimize for training time and computational constraints and were not selected for using any dataset.

### 7.5. Analysis Plan

All LLM-based approaches were designed to output a binary response (“yes” or “no”). Model outputs were parsed and mapped to integer labels: 1 for “yes” (stigmatizing), and 0 for “no” (non-stigmatizing). These labels were then used to compute evaluation metrics across the test dataset. Sample prompt shown in **Figure 4**.

We assessed performance using scikit-learn [18] accuracy and macro-averaged F1 score implementation, which accounts for performance across both classes irrespective of class imbalance. To estimate the variability of our metrics, we computed 95% confidence intervals via bootstrapping with 1,000 resamples. Two sample student t-test were performed on bootstrapped F1 macro scores to establish significant differences in scores.

Evaluation was conducted on four subsets of the test data:

1. Full Test Set – All clinical notes included in the held-out test split.
2. Subset with Potentially Stigmatizing Terms – Only notes that contained one or more stigmatizing terms as defined by the NIDA Guidelines [6].
3. External Validation Full from UW
4. External Validation Down-Sampled from UW

This second evaluation subset was included to assess model performance on the most challenging examples - those where potentially stigmatizing terms are present, but their contextual use may or may not be stigmatizing. This distinction is critical, as it tests the model’s ability to move beyond keyword detection to understand context and nuance in language.

The third and fourth subsets were datasets curated by UW and contained unseen progress notes by the approaches in training and/or hyperparameter selection phases. This was utilized to assess the generalizability of the approaches for the task of stigmatizing language detection. The full external validation set was utilized to assess how the approaches perform on detecting stigmatizing language at the native prevalence of such language. We also assessed the model on the down-sampled balanced subset to create a fair comparison between the MIMIC-derived dataset and external validation dataset, as this would allow us to understand if performance differences are solely due to the unseen nature of the external dataset.

#### 7.5.1. Error Analysis

To further characterize model behavior, we conducted a qualitative error analysis on the best-performing fine-tuned and best-performing non-fine-tuned models, as determined by validation set performance. Specifically, we focused on false positives: cases where the model incorrectly labeled non-stigmatizing notes as stigmatizing.

For each false positive instance, the model was prompted to explain its classification decision using a separate, explanatory prompt designed to elicit a natural language justification. This analysis served two purposes: (1) to evaluate the fluency and coherence of the model’s explanations, and (2) to determine whether the model’s errors reflected plausible misinterpretations or spurious correlations.

To maintain consistency with their original inference settings, the in-context learning configuration received the same contextual guidelines during explanation generation. The explanatory prompt used in this analysis is shown in **Figure 5**.

**Figure 5.**
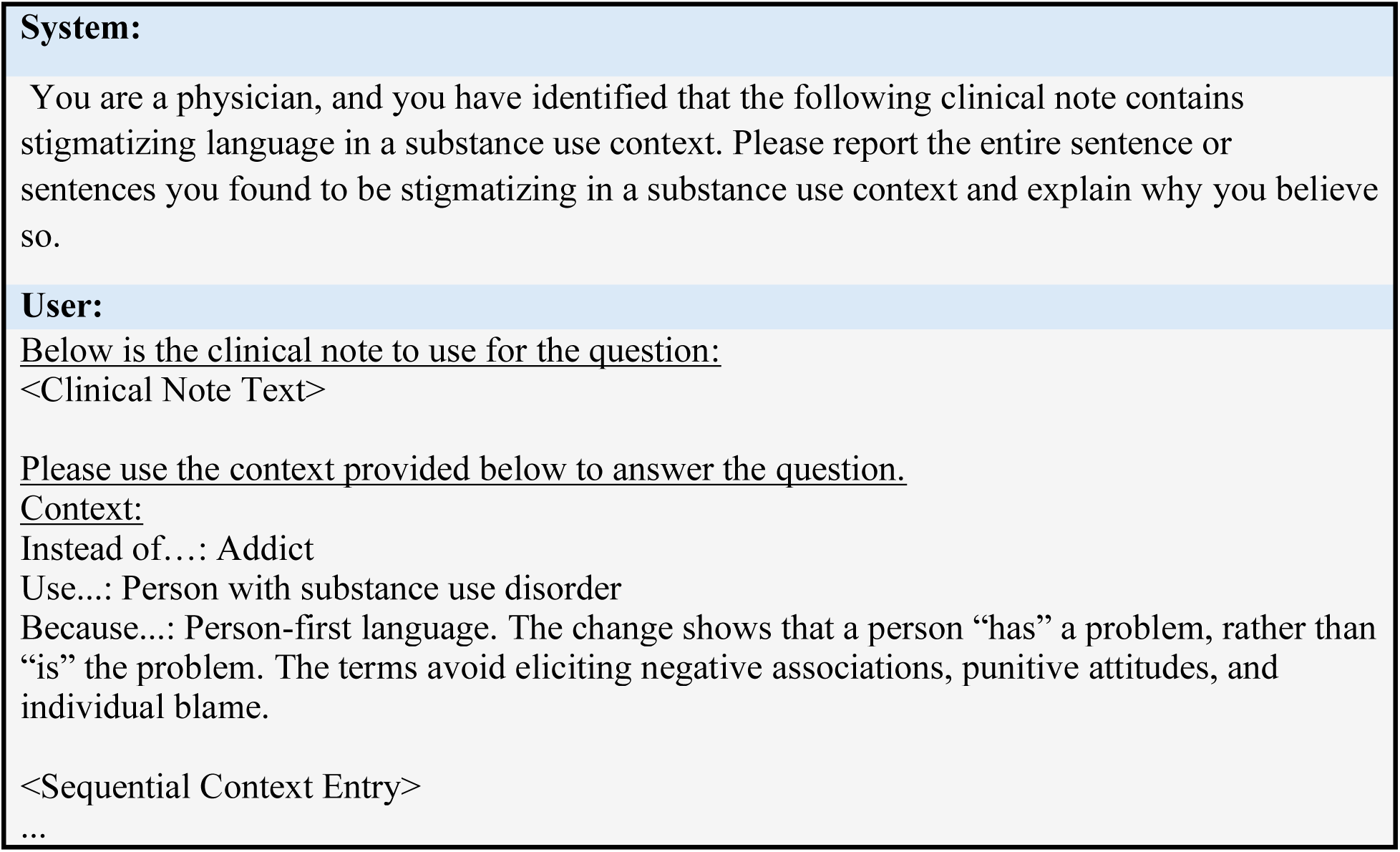
General Structure of Error Analysis Prompt. When applying this prompt to the RAG approach, there are n context entries. When applying the in-context approach, all available context entries are provided. Finally, when applying to the Zero-Shot or SFT approach, there is no context subfield in the user portion of the prompt. System prompts prime the LLM to answer user prompts following certain directions. Hence the system prompt gives the LLM a scenario and an output format.

This analysis provided insight into the types of linguistic cues the models considered stigmatizing and highlighted potential areas in which fine-tuning improved contextual understanding or exacerbated overfitting. The error analyses were evaluated and verified by a board-certified Addiction Medicine specialist and trainer on how to avoid stigmatizing language in addiction care (ESA).

## Acknowledgements

This work was funded by National Institutes of Health (NIH) R01LM012973, R01DA051464. The funder played no role in study design, data collection, analysis and interpretation of data, or the writing of this manuscript.

## Author Contributions

RS preprocessed train, validation, and test datasets. RS performed LLM training and inference, LLM approach engineering, prompt/context engineering, LLM evaluation (performance and error analysis). RS drafted the manuscript. JC engineered dataset labeling process and labeled train, validation, test, and external validation datasets. JC and AM ran external validation. ESA analyzed false positives for error analysis. DD and MA provided feedback on LLM approach engineering, evaluation, and manuscript editing. All authors read and reviewed the manuscript.

## Data Availability

The data that support the findings of this study are available from MIMIC [5] but restrictions apply to the availability of these data, which were used under license for the current study, and so are not publicly available. Data are however available from the authors upon reasonable request and with permission of MIMIC [5]. Data will be distributed through PhysioNet after the paper is accepted.

## Code Availability

The underlying code for this study is available in rsethi21/clinicallyStigmatizingLanguage and can be accessed via this link: https://github.com/rsethi21/clinicallyStigmatizingLanguage.

